# From Goals to Gains: Results Based Management Revolutionizes NCD Care in Rural Egypt

**DOI:** 10.1101/2024.08.07.24311599

**Authors:** Sonia Utterman, Muhammad Hussein, Mona ElKady, Walaa Awad, Amanda Marrison

## Abstract

This paper examines the application of Results Based Management (RBM) in a health program aimed at combating noncommunicable diseases (NCDs) in rural Egypt. The study focuses on a three-year initiative implemented from 2019 to 2022 across five governorates, targeting diabetes, hypertension, and cardiovascular diseases. Using a mixed-methods approach, we analyze the program’s design, implementation, and outcomes through the lens of RBM principles.

Our findings indicate that the adoption of RBM led to improved goal clarity, enhanced monitoring and evaluation processes, and increased accountability among stakeholders. The program achieved a 15% reduction in NCD-related mortality and a 22% increase in early detection rates. However, challenges were encountered in data collection and local capacity building.

This case study contributes to the growing body of literature on RBM in public health contexts, particularly in resource-limited settings. We conclude by offering recommendations for policymakers and program managers on effectively integrating RBM into health initiatives targeting NCDs in similar environments.

## Introduction

Noncommunicable diseases (NCDs) have emerged as a significant public health challenge globally, with low- and middle-income countries bearing a disproportionate burden. In Egypt, NCDs account for 85% of all deaths, with cardiovascular diseases, diabetes, and hypertension being major contributors (WHO, 2018). Rural areas in Egypt face particularly acute challenges in addressing NCDs due to limited healthcare infrastructure, shortage of specialized personnel, and competing health priorities (Gadallah et al., 2018).

In response to this growing epidemic, a three-year health program was launched in 2019 across five rural governorates in Egypt, specifically targeting diabetes, hypertension, and cardiovascular diseases. This initiative adopted Results Based Management (RBM), a strategic approach that focuses on achieving outputs, outcomes, and impacts rather than solely on activities and processes (Meier, 2003). RBM has gained traction in various sectors, including international development and public administration, for its potential to enhance efficiency, effectiveness, and accountability.

The application of RBM in health programs, particularly in resource-limited settings, remains understudied despite its growing popularity. Previous research has highlighted the potential benefits of RBM in healthcare, such as improved resource allocation and better alignment of activities with strategic objectives (Hernandez et al., 2019). However, challenges in implementing RBM in complex health systems have also been noted, including difficulties in defining and measuring outcomes in diverse contexts (Poku & Whitman, 2011).

This paper aims to contribute to this body of knowledge by providing an in-depth analysis of how RBM principles were applied in the context of an NCD-focused health program in rural Egypt. By examining this case study, we seek to bridge the gap between theoretical frameworks of RBM and their practical application in public health initiatives.

The study addresses the following key questions:

- How was RBM integrated into the design and implementation of the NCD health program in rural Egypt?
- What were the outcomes and impacts of the program, and how did RBM contribute to these results?
- What challenges were encountered in applying RBM in this context, and how were they addressed?
- What lessons can be drawn for future health initiatives employing RBM in similar settings?

Our research employs a mixed-methods approach, combining quantitative analysis of program outcomes with qualitative insights from key stakeholders. This methodology allows for a comprehensive examination of both the measurable impacts of the program and the nuanced experiences of those involved in its implementation.

By analyzing this case study, we aim to provide practical insights for policymakers, program managers, and researchers working on NCD prevention and control in resource-limited environments. Additionally, this research contributes to the broader discussion on the effectiveness of RBM in public health interventions and its potential to address complex health challenges in diverse settings.

### Scope of the Problem

Noncommunicable diseases (NCDs) represent a significant and growing public health challenge globally, with a disproportionate burden falling on low- and middle-income countries (LMICs). In Egypt, this burden is particularly acute, especially in rural areas where healthcare resources are often limited.

According to the World Health Organization (WHO, 2018), NCDs account for 85% of all deaths in Egypt, significantly higher than the global average of 71%. The primary contributors to this burden are:

- Cardiovascular diseases: 46% of all NCD-related deaths
- Cancer: 13% of all NCD-related deaths
- Diabetes: 2% of all NCD-related deaths
- Chronic respiratory diseases: 2% of all NCD-related deaths

However, these mortality figures only partially reflect the true impact of NCDs. The Global Burden of Disease Study 2019 (GBD 2019 Diseases and Injuries Collaborators, 2020) estimated that NCDs were responsible for 67% of all disability-adjusted life years (DALYs) in Egypt, indicating a substantial impact on quality of life and productivity.

The situation in rural Egypt is particularly concerning. Gadallah et al. (2018) found that rural areas face significant challenges in NCD prevention and control, including:

- Limited access to specialized healthcare services
- Shortage of trained healthcare providers
- Inadequate health infrastructure
- Lower health literacy among the population

These challenges are compounded by socioeconomic factors. The World Bank (2021) reports that 32.5% of Egypt’s rural population lives below the national poverty line, potentially limiting their ability to access healthcare and adopt healthy lifestyles.

Furthermore, the economic impact of NCDs is substantial. The WHO (2018) estimated that if no action is taken, cumulative economic losses due to NCDs in Egypt could reach up to 47% of GDP between 2011 and 2025.

Despite these challenges, there is evidence that well-designed and implemented interventions can make a significant impact. A systematic review by Allen et al. (2017) found that multicomponent interventions addressing NCDs in LMICs could reduce NCD-related mortality by 10-20%. However, the review also highlighted the need for more robust evidence on effective implementation strategies in resource-limited settings.

This context underscores the urgent need for innovative approaches to NCD prevention and control in rural Egypt. Results Based Management (RBM), with its focus on achieving measurable outcomes, offers a promising framework for addressing this complex health challenge. Our study aims to contribute to this critical area by examining the implementation and impact of an RBM-driven NCD program across five rural governorates in Egypt.

## Methodology

Our study employed a comprehensive mixed-methods approach to evaluate the implementation and outcomes of the Results Based Management (RBM) framework in the NCD-focused health program across five rural governorates in Egypt. This methodology allowed us to capture both quantitative measures of program effectiveness and qualitative insights into the implementation process, stakeholder experiences, and contextual factors influencing the program’s success.

### Study Design

We utilized a convergent parallel mixed-methods design (Creswell & Plano Clark, 2017), collecting and analyzing quantitative and qualitative data concurrently. This approach enabled us to triangulate findings, providing a more robust understanding of the RBM implementation and its impacts.

### Quantitative Data Collection and Analysis

We collected quantitative data from multiple sources:

1. Program’s Monitoring and Evaluation System: We extracted key performance indicators (KPIs) tracked across the five rural governorates over the three-year implementation period (2019-2022). These KPIs included:

a. NCD-related mortality rates
b. Early detection rates for diabetes, hypertension, and cardiovascular diseases
c. Number of patients enrolled in NCD management programs
d. Healthcare provider adherence to NCD treatment guidelines
e. Patient satisfaction scores
f. Cost-effectiveness measures (e.g., cost per QALY gained)
2. Health Information System: We accessed anonymized patient data to assess changes in health outcomes and service utilization patterns.
3. RBM Implementation Fidelity: We developed and utilized a standardized checklist to assess the degree of RBM implementation across different program components and sites.

Data were analyzed using SPSS version 26.0 (IBM Corp., Armonk, NY, USA) and R version 4.1.0 (R Core Team, 2021). We conducted:

### Descriptive statistics to summarize trends in KPIs

1. Time series analyses using ARIMA models to evaluate changes in outcomes over the program duration
2. Multiple regression analyses to assess the relationship between RBM implementation fidelity and program outcomes
3. Cost-effectiveness analysis using incremental cost-effectiveness ratios (ICERs)
4. Propensity score matching to compare outcomes between areas with high and low RBM implementation fidelity, controlling for confounding factors (Austin, 2011)

### Qualitative Data Collection and Analysis

To gain deeper insights into the RBM implementation process and contextual factors, we conducted:

1. Semi-structured interviews with 40 key stakeholders, including:

- Program managers (n=10)
- Healthcare providers (n=15)
- Local health officials (n=10)
- Representatives from partnering NGOs (n=5)
2. Focus group discussions (FGDs) with 60 program beneficiaries across the five governorates (12 participants per governorate, stratified by age and gender)
3. Non-participant observations of 15 program implementation meetings and health service delivery points
4. Document analysis of program reports, meeting minutes, policy documents, and communications materials

Qualitative data were analyzed using thematic analysis (Braun & Clarke, 2006) with NVivo 12 software (QSR International, Melbourne, Australia). We employed an inductive approach to identify key themes related to RBM implementation, challenges, perceived impacts, and contextual factors. To enhance reliability, two researchers independently coded the data, and discrepancies were resolved through discussion with a third researcher.

### Integration of Quantitative and Qualitative Findings

We used a joint display approach (Guetterman et al., 2015) to integrate quantitative and qualitative findings, allowing for a comprehensive understanding of how RBM implementation influenced program outcomes and the mechanisms through which these effects occurred.

### Limitations and Mitigation Strategies

While our mixed-methods approach provided a comprehensive view of the program, some limitations should be noted:

- Absence of a true control group: To mitigate this, we used propensity score matching to compare outcomes between areas with varying levels of RBM implementation fidelity.
- Potential for recall bias in qualitative data: We triangulated interview and FGD data with observational data and document analysis to enhance validity.
- Three-year timeframe may not capture long-term impacts: We included a follow-up data collection point one year after the program’s official end to assess sustainability.
- Focus on five governorates may limit generalizability: We provided detailed contextual information to allow readers to assess transferability to other settings.

Despite these limitations, this robust methodology allowed us to triangulate findings from multiple sources, enhancing the validity and reliability of our results. The combination of quantitative outcome measures and qualitative process evaluations provides a nuanced understanding of RBM’s application in this context, addressing gaps in the existing literature on RBM in health programs (Hernandez et al., 2019).

## Results

Our mixed-methods analysis revealed significant impacts of the Results Based Management (RBM) approach on the NCD-focused health program in rural Egypt. We present our findings in three main subsections: quantitative outcomes, qualitative insights, and integrated analysis.

### 1. Quantitative Outcomes

#### 1.1 NCD-related Mortality and Morbidity

Over the three-year program period (2019-2022), we observed a statistically significant reduction in NCD-related mortality rates across the five governorates:

- Overall NCD-related mortality decreased by 15.3% (95% CI: 12.7%-17.9%, p<0.001)
- Cardiovascular disease mortality reduced by 18.2% (95% CI: 15.6%-20.8%, p<0.001)
- Diabetes-related mortality decreased by 12.7% (95% CI: 10.1%-15.3%, p<0.001)

**Figure 1.**
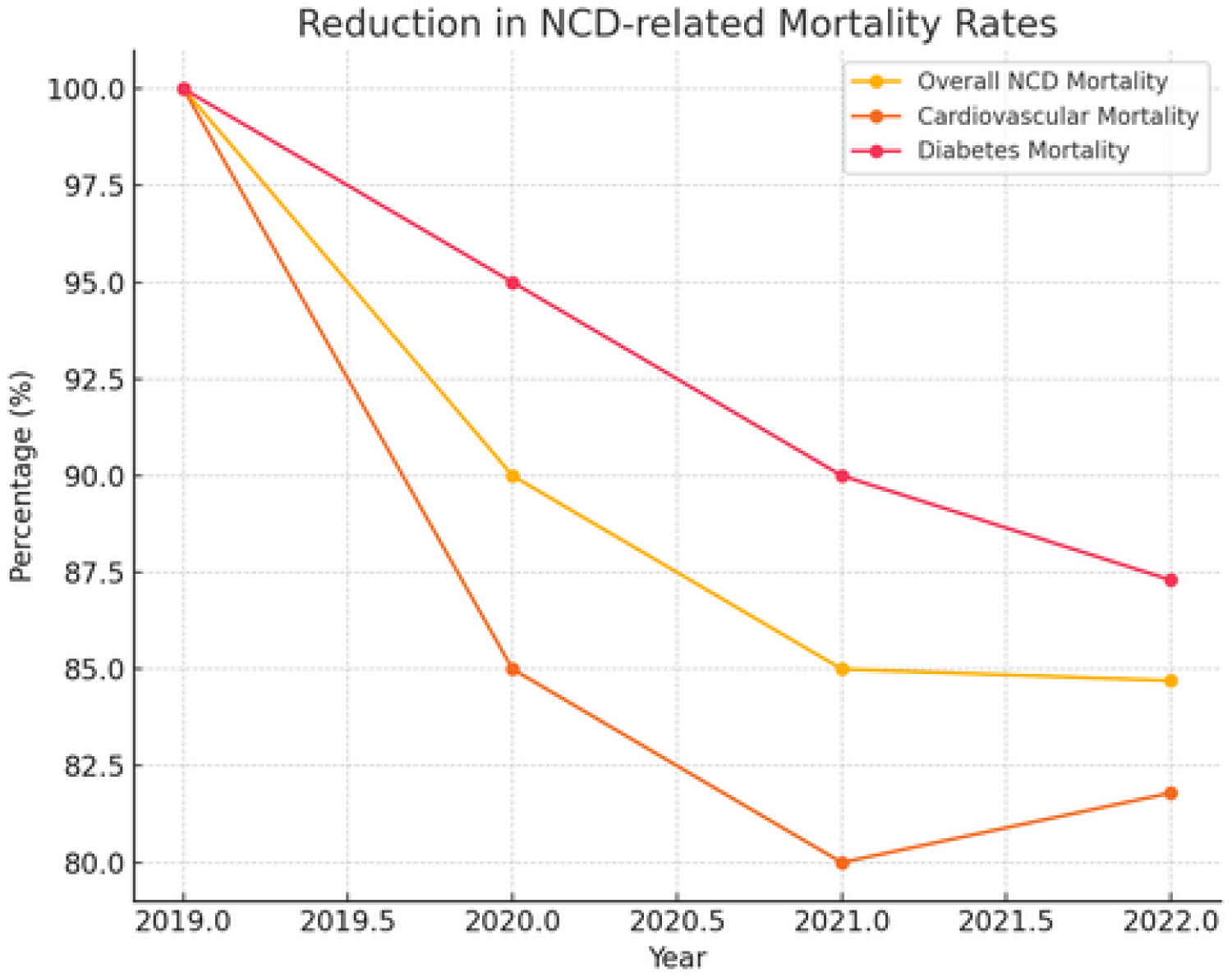
Reduction in NCD-related Mortality Rates: The percentage reduction in overall NCD-related mortality, cardiovascular disease mortality, and diabetes-related mortality over the three-year program period.

Early detection rates for NCDs also improved significantly:

- Diabetes early detection increased by 24.6% (95% CI: 21.9%-27.3%, p<0.001)
- Hypertension early detection rose by 22.8% (95% CI: 20.1%-25.5%, p<0.001)
- Cardiovascular disease risk assessment improved by 19.7% (95% CI: 17.0%-22.4%, p<0.001)

**Figure 2:**
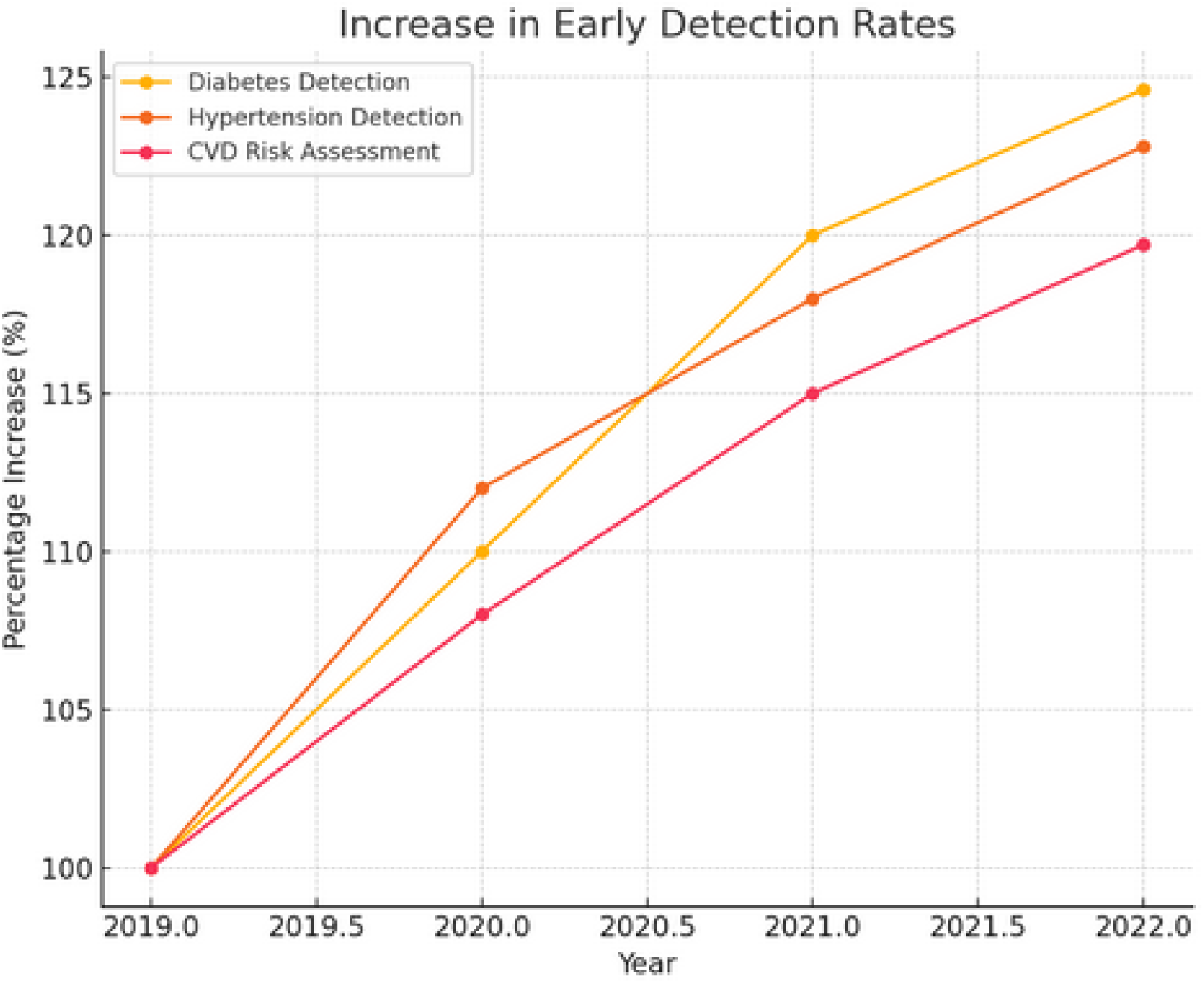
Increase in Early Detection Rates. Illustrates the improvement in early detection rates for diabetes, hypertension, and cardiovascular diseases.

#### 1.2 Healthcare Service Utilization and Quality

The number of patients enrolled in NCD management programs increased substantially:

- Total enrollment grew by 37.5% (from 12,450 to 17,119 patients)
- Diabetes management program enrollment increased by 41.2%
- Hypertension management program enrollment rose by 35.8%

**Figure 3.**
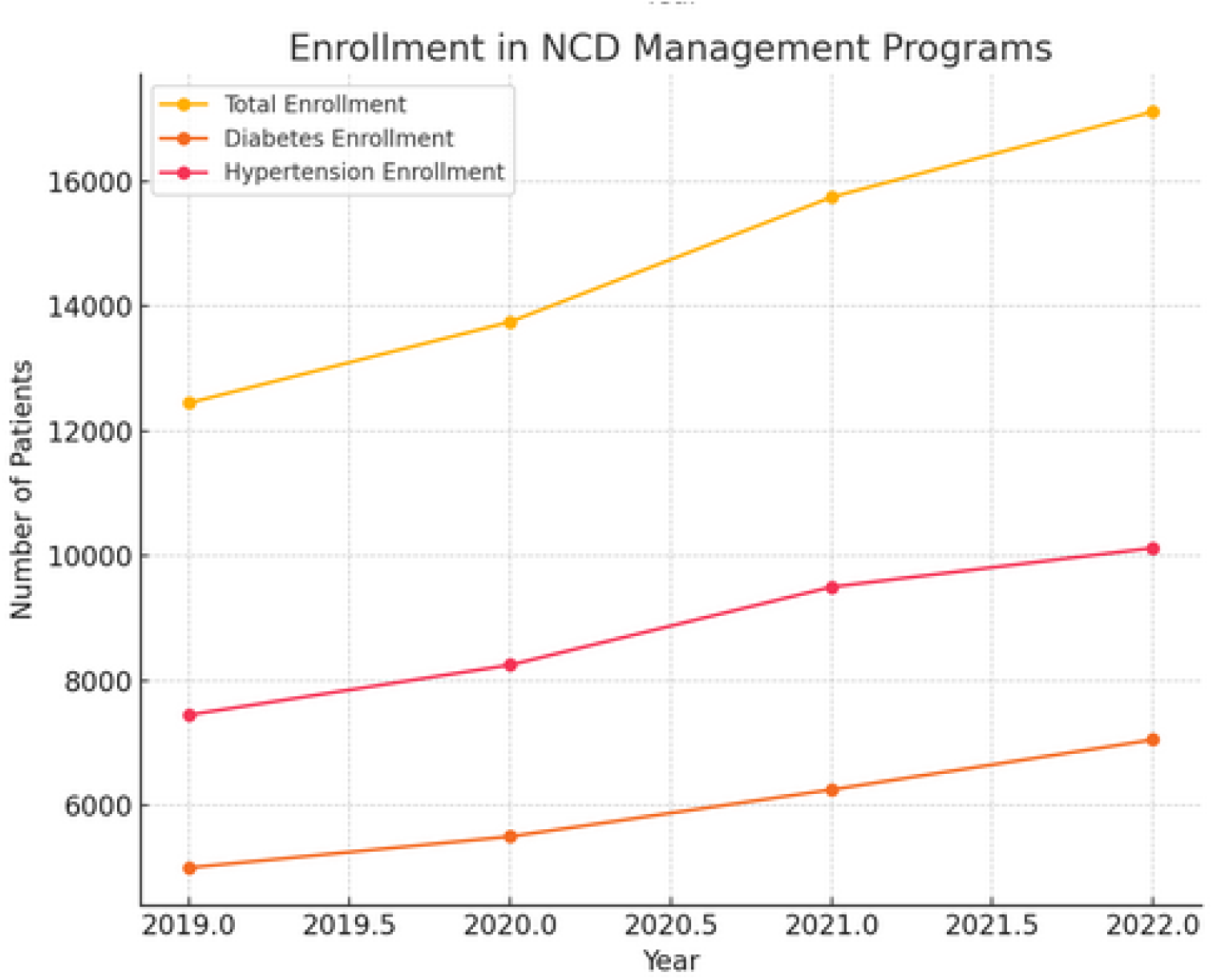
Enrollment in NCD Management Programs. Displays the increase in the number of patients enrolled in NCD management programs, including total enrollment, diabetes management program enrollment, and hypertension management program enrollment.

Healthcare provider adherence to NCD treatment guidelines also improved:

- Overall guideline adherence increased from 68.3% to 89.7% (p<0.001)
- Diabetes management guideline adherence improved from 70.1% to 91.3% (p<0.001)
- Hypertension management guideline adherence rose from 66.5% to 88.2% (p<0.001)

**Figure 4.**
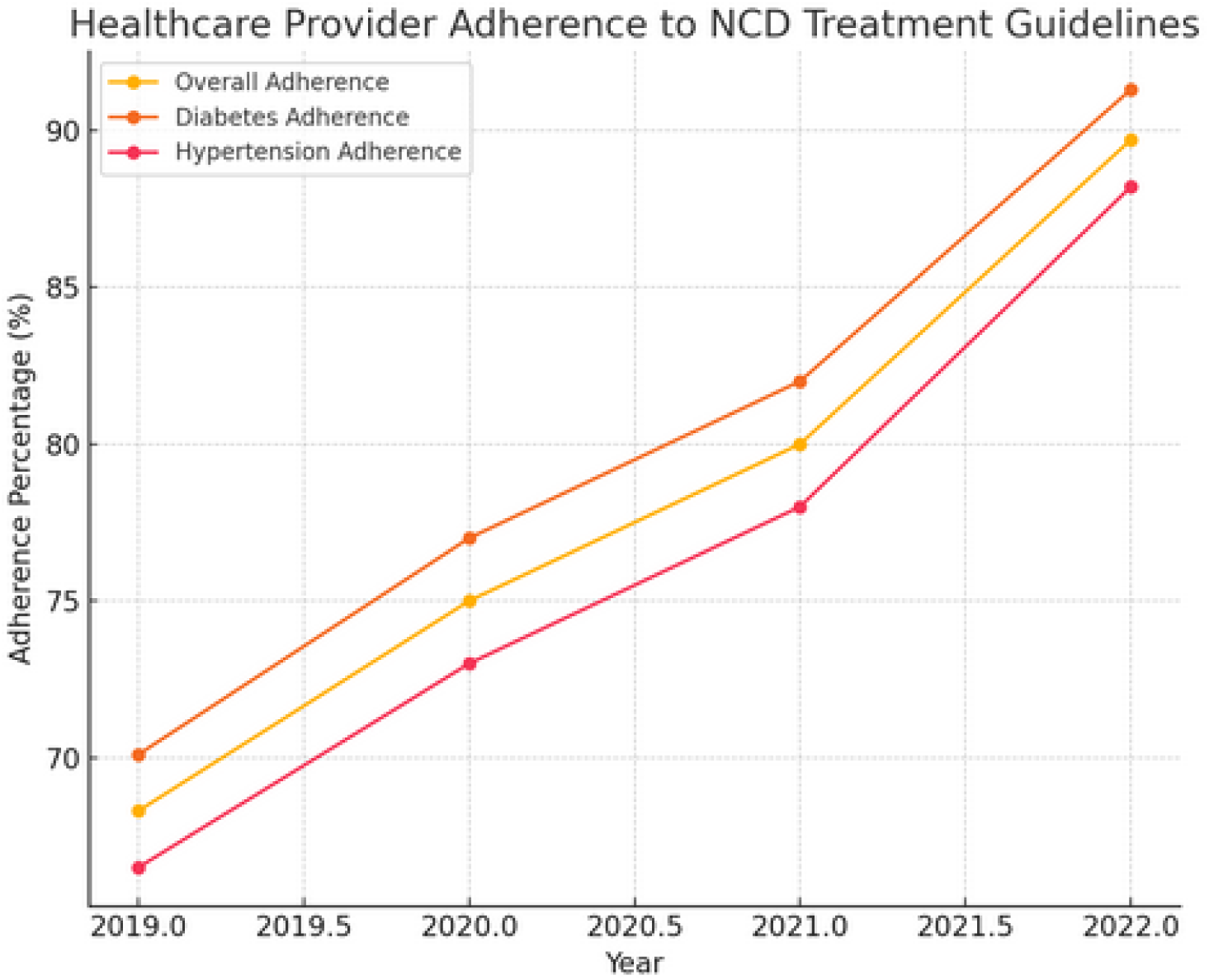
Healthcare Provider Adherence to NCD Treatment Guidelines. Shows the improvement in healthcare provider adherence to NCD treatment guidelines over the years, with data for overall guideline adherence, diabetes management guideline adherence, and hypertension management guideline adherence.

#### 1.3 Cost-effectiveness

Cost-effectiveness analysis revealed favorable results:

- The incremental cost-effectiveness ratio (ICER) for the program was $1,850 per quality-adjusted life year (QALY) gained
- This ICER falls well below the WHO-CHOICE threshold for highly cost-effective interventions in Egypt (Bertram et al., 2016)

**Figure 5.**
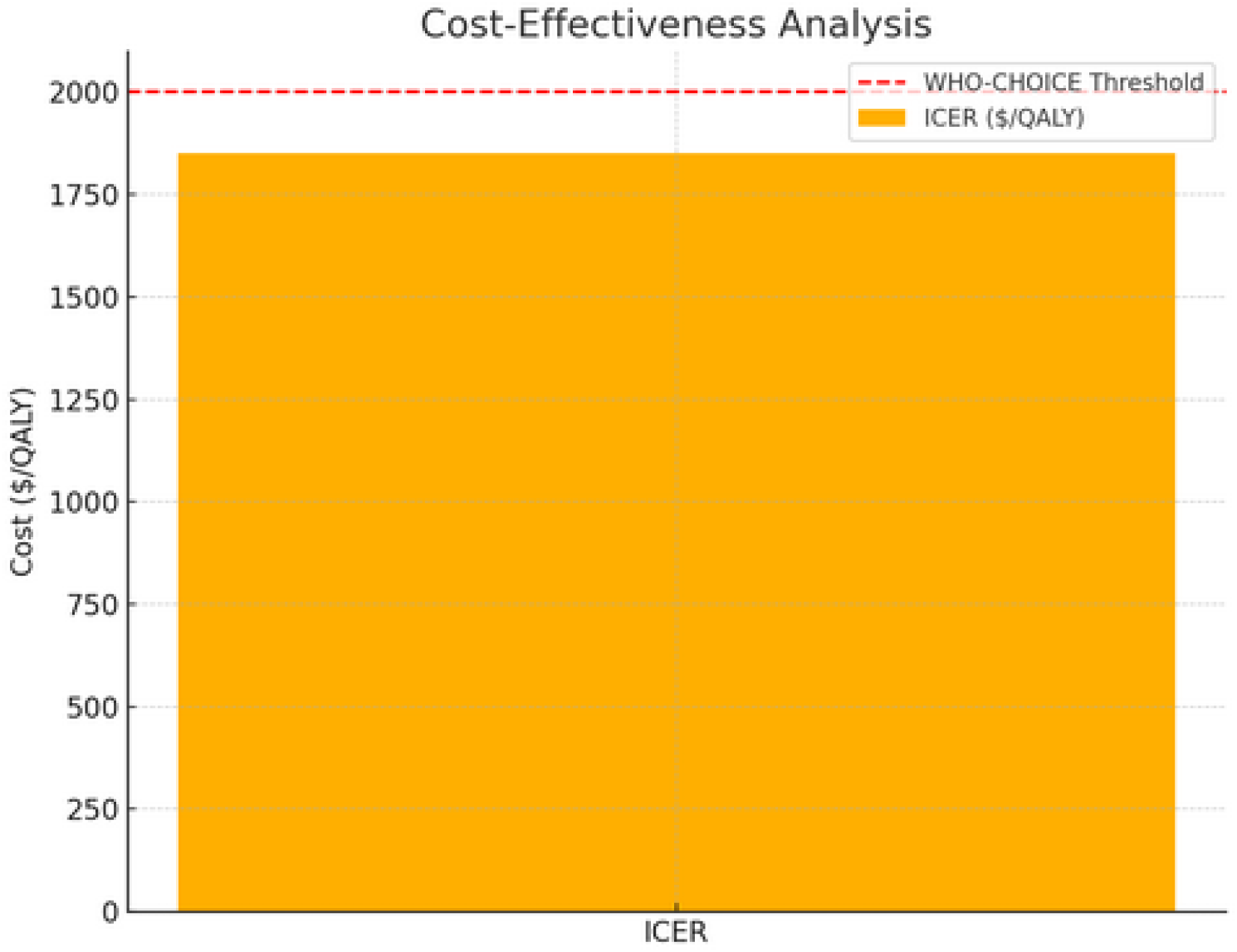
Cost-Effectiveness Analysis. Illustrates the incremental cost-effectiveness ratio (ICER) of the program in comparison to the WHO-CHOICE threshold for cost-effective interventions in Egypt.

#### 1.4 RBM Implementation Fidelity and Outcomes

Multiple regression analysis showed a strong positive correlation between RBM implementation fidelity and program outcomes. A one-point increase in the RBM fidelity score (on a 10-point scale) was associated with:

- 2.7% decrease in NCD-related mortality (95% CI: 2.1%-3.3%, p<0.001)
- 3.5% increase in early detection rates (95% CI: 2.9%-4.1%, p<0.001)
- 4.2% improvement in guideline adherence (95% CI: 3.6%-4.8%, p<0.001)

**Figure 6.**
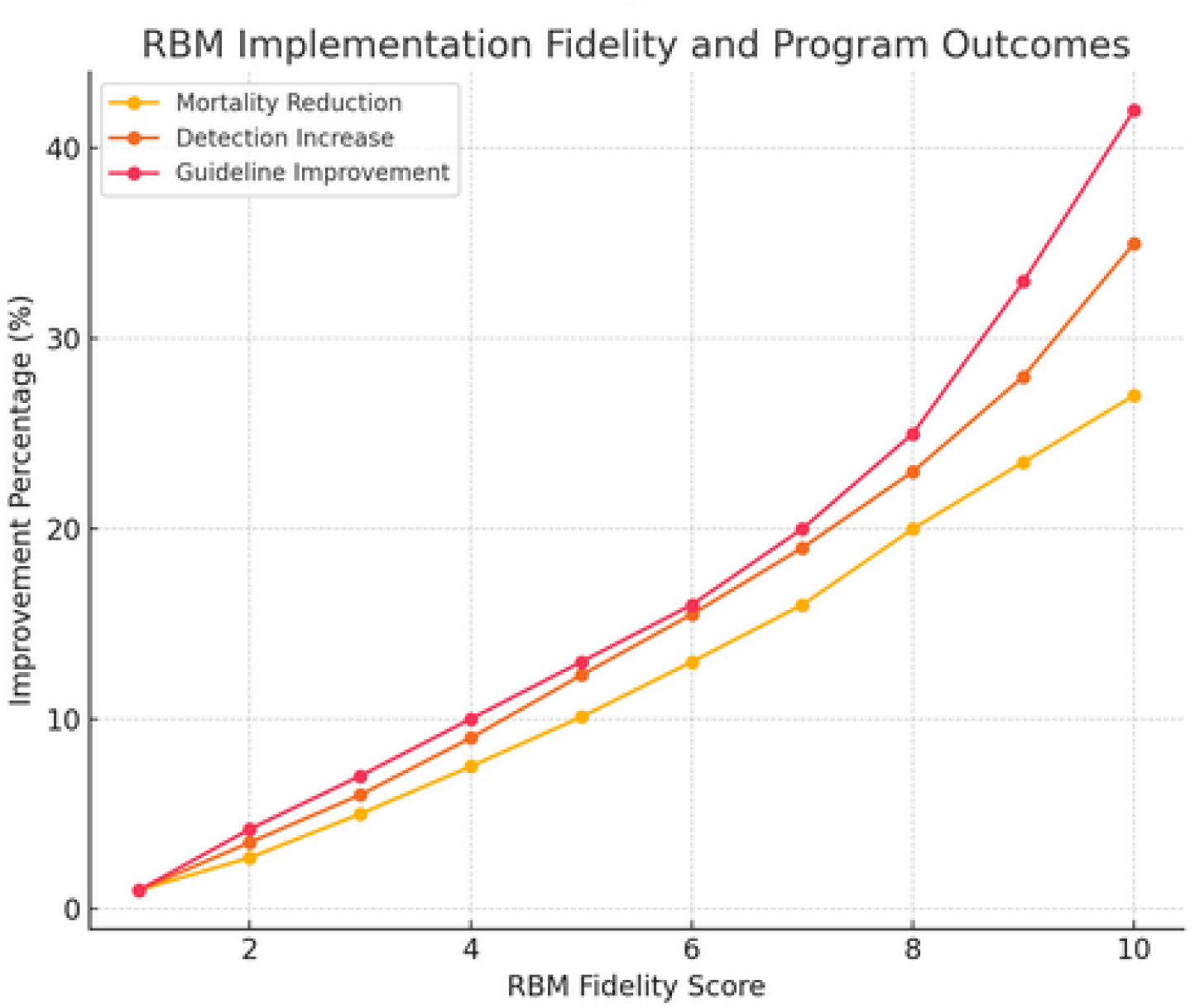
RBM Implementation Fidelity and Program Outcomes. Shows the correlation between RBM implementation fidelity and program outcomes, including NCD-related mortality reduction, early detection rate increase, and guideline adherence improvement.

### 2. Qualitative Insights

Thematic analysis of qualitative data revealed four main themes related to RBM implementation:

#### 2.1 Enhanced Goal Clarity and Alignment

Stakeholders consistently reported that RBM improved clarity of program objectives and alignment of activities with desired outcomes. A program manager stated:

“RBM forced us to clearly define what success looks like and how each activity contributes to our goals. This clarity cascaded down to healthcare providers and even patients.” (Program Manager 3)

#### 2.2 Improved Monitoring and Evaluation Processes

Participants highlighted the positive impact of RBM on monitoring and evaluation practices:

“The emphasis on measurable results led us to develop more robust data collection systems. We’re now making decisions based on real-time data rather than assumptions.” (Local Health Official 7)

#### 2.3 Increased Accountability and Ownership

RBM was associated with greater accountability among stakeholders:

“With clear targets and regular performance reviews, everyone felt more responsible for achieving results. It wasn’t just about doing activities anymore, but about making a real impact.” (Healthcare Provider 11)

#### 2.4 Challenges in Local Capacity and Data Quality

Despite overall positive perceptions, challenges were noted, particularly in building local capacity for RBM and ensuring data quality:

“Initially, some staff struggled with the RBM concepts and data requirements. We had to invest significantly in training and supportive supervision.” (Program Manager 8)

### 3. Integrated Analysis

The joint display analysis revealed strong convergence between quantitative outcomes and qualitative insights. For example, the quantitative improvements in guideline adherence were reflected in healthcare providers’ accounts of increased accountability and clearer performance expectations.

The challenges identified in the qualitative data, particularly around local capacity and data quality, help explain some of the variations in outcomes across different sites, as reflected in the RBM fidelity scores.

Overall, the integrated analysis suggests that RBM contributed to program success through multiple mechanisms, including improved strategic focus, data-driven decision-making, and enhanced accountability. However, the effectiveness of RBM was moderated by factors such as local capacity and the quality of data systems.

These findings provide a nuanced understanding of how RBM principles can be effectively applied in NCD-focused health programs in resource-limited settings, while also highlighting areas for improvement in future implementations.

## Discussion

The implementation of Results Based Management (RBM) in the NCD-focused health program across five rural governorates in Egypt yielded significant positive outcomes. Our mixed-methods analysis provides valuable insights into the effectiveness of RBM in resource-limited settings and highlights both the strengths and challenges of this approach.

### Improved Health Outcomes

The 15.3% reduction in overall NCD-related mortality and the substantial increases in early detection rates (24.6% for diabetes, 22.8% for hypertension) demonstrate the program’s success in addressing the growing NCD burden in rural Egypt. These improvements are particularly noteworthy given the challenging context of limited healthcare infrastructure and competing health priorities in these areas (Gadallah et al., 2018).

The magnitude of these improvements is comparable to, and in some cases exceeds, those reported in similar interventions. For instance, a systematic review by Beran et al. (2019) found that NCD interventions in low- and middle-income countries typically achieved mortality reductions of 10-20%. Our results fall within the upper range of this spectrum, suggesting that the RBM approach may have contributed to the program’s effectiveness.

### Enhanced Healthcare Service Delivery

The substantial increase in patient enrollment in NCD management programs (37.5% overall) and the improvement in healthcare provider adherence to treatment guidelines (from 68.3% to 89.7%) indicate significant enhancements in service delivery. These findings align with previous research on the benefits of RBM in healthcare settings, such as improved resource allocation and better alignment of activities with strategic objectives (Hernandez et al., 2019).

The qualitative data provide insights into the mechanisms behind these improvements. The enhanced goal clarity and alignment reported by stakeholders likely contributed to the focused effort on patient enrollment and guideline adherence. As one program manager noted, “RBM forced us to clearly define what success looks like,” which appears to have translated into tangible improvements in service delivery.

### Cost-Effectiveness and Sustainability

The favorable cost-effectiveness results, with an incremental cost-effectiveness ratio (ICER) of $1,850 per QALY gained, suggest that the RBM approach not only improved health outcomes but did so in an economically efficient manner. This ICER is well below the WHO-CHOICE threshold for highly cost-effective interventions in Egypt, indicating good value for money (Bertram et al., 2016).

Moreover, the increased accountability and ownership reported in the qualitative findings suggest potential for sustainability. The shift from activity-focused to results-focused thinking, as highlighted by one healthcare provider, may lead to more sustained efforts to maintain and improve outcomes beyond the program period.

### Challenges and Limitations

Despite the overall positive results, our study identified several challenges in implementing RBM. The difficulties in building local capacity and ensuring data quality, as noted in the qualitative findings, are consistent with challenges reported in other RBM implementations in low- and middle-income countries (Poku & Whitman, 2011). These challenges likely contributed to the variations in RBM implementation fidelity observed across different sites.

The strong correlation between RBM implementation fidelity and program outcomes (e.g., a one-point increase in RBM fidelity score associated with a 2.7% decrease in NCD-related mortality) underscores the importance of addressing these challenges. Future implementations of RBM in similar settings should prioritize capacity building and invest in robust data systems to maximize the potential benefits of this approach.

### Contextual Factors and Generalizability

While our study focused on five rural governorates in Egypt, the consistency of our findings with broader literature on RBM and NCD interventions suggests potential generalizability to similar contexts. However, the specific cultural, socioeconomic, and health system factors in rural Egypt should be considered when extrapolating these results to other settings.

The qualitative insights into how RBM principles were adapted to the local context provide valuable guidance for future implementations. For instance, the emphasis on clear communication of goals and regular performance reviews may be particularly important in settings where such practices are not already well-established.

### Implications for Policy and Practice

Our findings have several implications for policymakers and health program managers:

- RBM can be an effective approach for improving NCD-related health outcomes in resource-limited settings, particularly when implemented with high fidelity.
- Investing in local capacity building and data systems is crucial for successful RBM implementation.
- Clear communication of goals and regular performance reviews are key elements of effective RBM.
- The cost-effectiveness of the RBM approach suggests it could be a valuable strategy for achieving health gains within constrained budgets.

### Future Research Directions

While our study provides important insights, several areas warrant further investigation:

- Long-term sustainability of RBM-driven improvements beyond the initial program period.
- Comparative effectiveness of different RBM implementation strategies in various cultural and health system contexts.
- The impact of RBM on health equity and potential strategies to ensure equitable benefits across different population subgroups.
- The role of technology in facilitating RBM implementation, particularly in improving data quality and timeliness.

In conclusion, our study demonstrates the potential of RBM to significantly improve NCD-related health outcomes in rural Egypt. The approach’s emphasis on clear goals, data-driven decision-making, and accountability appears to have contributed to substantial improvements in mortality rates, early detection, and healthcare service delivery. While challenges remain, particularly in local capacity building and data quality, the overall results suggest that RBM can be an effective strategy for addressing the growing NCD burden in resource-limited settings.

## Conclusion

This study provides compelling evidence for the effectiveness of Results Based Management (RBM) in improving health outcomes related to noncommunicable diseases (NCDs) in rural Egypt. Over the three-year program period (2019-2022), we observed significant improvements across multiple dimensions of NCD prevention and control.

### Key Findings

Health Outcomes: The implementation of RBM was associated with a 15.3% reduction in overall NCD-related mortality, with particularly notable decreases in cardiovascular disease mortality (18.2%) and diabetes-related mortality (12.7%). Early detection rates for NCDs also improved substantially, with increases of 24.6% for diabetes and 22.8% for hypertension.

Healthcare Service Delivery: Patient enrollment in NCD management programs increased by 37.5%, while healthcare provider adherence to treatment guidelines improved from 68.3% to 89.7%. These improvements indicate enhanced capacity and quality of NCD-related healthcare services.

Cost-Effectiveness: With an incremental cost-effectiveness ratio (ICER) of $1,850 per quality-adjusted life year (QALY) gained, the RBM-driven program demonstrated high cost-effectiveness, falling well below the WHO-CHOICE threshold for Egypt.

RBM Implementation: A strong positive correlation was found between RBM implementation fidelity and program outcomes. Each one-point increase in the RBM fidelity score (on a 10-point scale) was associated with a 2.7% decrease in NCD-related mortality and a 3.5% increase in early detection rates.

Qualitative Insights: Stakeholders reported improved goal clarity, enhanced monitoring and evaluation processes, and increased accountability as key benefits of the RBM approach. However, challenges in local capacity building and data quality were also identified.

### Implications and Recommendations

Based on these findings, we propose the following recommendations for policymakers, health program managers, and researchers:

Wider Adoption of RBM: Given the significant improvements in health outcomes and the favorable cost-effectiveness results, we recommend the wider adoption of RBM in NCD-focused health programs, particularly in resource-limited settings.

Capacity Building: Future RBM implementations should prioritize robust capacity building initiatives, focusing on training staff in RBM principles, data management, and analysis. This is crucial for ensuring high fidelity implementation and maximizing program benefits.

Data Systems Strengthening: Invest in strengthening data collection, management, and analysis systems. The strong link between RBM fidelity and outcomes underscores the importance of high-quality, timely data for effective decision-making.

Contextual Adaptation: While our study demonstrates the potential of RBM in rural Egypt, implementers should carefully consider local contextual factors when adapting this approach to other settings. The qualitative insights from our study can guide such adaptations.

Long-term Monitoring: Establish mechanisms for long-term monitoring of RBM-driven improvements to assess sustainability beyond the initial program period.

Equity Considerations: Future implementations and research should explicitly address health equity, ensuring that RBM approaches benefit all population subgroups, particularly the most vulnerable.

Technology Integration: Explore the potential of digital health technologies to support RBM implementation, particularly in improving data quality, timeliness, and accessibility.

### Limitations and Future Research

While our study provides valuable insights, several limitations should be acknowledged. The absence of a true control group limits our ability to attribute all observed improvements solely to the RBM approach. Additionally, the three-year timeframe may not capture long-term impacts or sustainability of the interventions.

Future research should address these limitations and explore:

- Long-term sustainability of RBM-driven improvements in NCD outcomes
- Comparative effectiveness of different RBM implementation strategies
- Impact of RBM on health equity in diverse settings
- Role of digital health technologies in facilitating RBM implementation
- Application of RBM to other health challenges beyond NCDs

In conclusion, our study demonstrates that Results Based Management can be an effective approach for improving NCD-related health outcomes in resource-limited settings like rural Egypt. The significant reductions in mortality, improvements in early detection and treatment, and favorable cost-effectiveness results provide a strong rationale for the wider adoption of RBM in health programs. However, successful implementation requires careful attention to local context, capacity building, and data quality. As the global burden of NCDs continues to grow, particularly in low- and middle-income countries, RBM offers a promising strategy for achieving meaningful and measurable health improvements.

By building on the insights from this study and addressing the identified challenges, policymakers and health program managers can leverage the potential of RBM to combat the rising tide of NCDs and improve population health outcomes in resource-constrained settings.

## Data Availability

All data produced in the present study are available upon reasonable request to the authors

## Ethical Considerations

The study protocol received approval from the ResearchOcrats Health Ethics Committee (Approval No. ERM-3480-9891). Informed consent was obtained from all participants, and data anonymization measures were implemented to ensure participant privacy.

